# Analysis of 193,618 trauma patient presentations in war-affected Syria from July 2013 to July 2015

**DOI:** 10.1101/2024.01.10.21255221

**Authors:** Hani Mowafi, Mahmoud Hariri, Baobao Zhang, Houssam Alnahhas, Basil Bakri, Adam Eldahan, Moustafa Moustafa, Maher Saqqur, Anas Al-Kassem

**Author notes:** Corresponding Author: Hani Mowafi, MD, MPH, Associate Professor of Emergency Medicine, Department of Emergency Medicine, Yale University School of Medicine 464 Congress Ave, Suite 260, New Haven, CT 06519-1315. These authors contributed equally to the work (Hani Mowafi, Mahmoud Hariri).

## Abstract

**Introduction:** Since 2011, the Syrian war has produced a mounting toll in terms of deaths and displaced persons. We present an analysis of demographic and temporal patterns of trauma patient presentations to Syrian hospitals in non-governmental, non-Islamic State (NGNI) regions from 2013 – 2015.

**Methods:** We analyzed an administrative dataset of patient presentations to 95 NGNI Syrian hospitals in regions outside of Syrian government control from July 2013 – July 2015. Descriptive analysis of this secondary data is reported and logistic regression was performed to assess for factors associated with inpatient mortality.

**Results:** 193,618 trauma patients presented to 95 NGNI hospitals from July 2013 – July 2015 (154,225 male, 79.7%; 39,393 female, 20.4%). Age information was complete for 160,237 encounters (82.8%): 0-2y: 8,257 (4.3%), 3-12y: 24,199 (12.5%), 13-18y: 22,482 (11.6%), 19-60y: 100,553 (51.9%), and elders over 60 years: 4,746 (2.5%). 59,387 patients were admitted (Ward 57,625; ICU 1,762) for an average length of stay of 3.80 days. There were 2,694 inpatient deaths (4.5% of admitted) and 4,758 patients (8.0%) required transfer to another facility for definitive care. Shrapnel (81,946; 42.3%) and blunt/crush injuries (71,477; 36.9%) were dominant injury mechanisms with an increasing proportion of these injuries over time. Inpatient mortality was most associated with extremes of age (age less than 2 aOR 2.92; age greater than 60 aOR 2.48), penetrating chest trauma (gunshot-chest aOR 6.03) and neurotrauma (blast-head aOR 13.42; blast-spine aOR 11.31; gunshot-head aOR 10.07; shrapnel-head aOR 6.34). Civilians presentations increased from 20% at start of data collection to a peak of 50% in June 2015.

**Conclusion:** The Syrian war has resulted in large volumes of trauma patients and significant mortality at NGNI Syrian hospitals. Mortality was most associated with neurotrauma and penetrating chest trauma. There was an increasing trend over time towards blunt/crush and shrapnel injuries consistent with the transition to the widespread use of aerial bombardment with resultant explosions and building collapse. Civilians including children and the elderly represent high proportions of the injured in NGNI Syrian hospitals. Additional work is needed to improve documentation of clinical service and to assess outcomes of care to improve quality of services provided to Syrian war trauma patients.

## Introduction

As the world marks the 10^th^ year since the onset of the Syrian War, there is still much to learn about the health impact of the conflict inside Syria, on those displaced from the country and on the neighboring countries who have absorbed the majority of refugeese. The war has produced one of the largest humanitarian crises in recent memory with up to 570,000 Syrians killed(1) (the Unite Nations stopped recording after 100,000 verified through July 2013(2)) with more than 1.5 million civilians seriously wounded.(3) Armed clashes and aerial bombardment of population areas produced the largest global displacement due to conflict,(4) with an estimated 6.7 million internally displaced persons (IDPs) and over 6.6 million refugees – 5.6 million of whom are in neighboring countries.(5, 6) The influx of refugees into the neighboring countries of Lebanon, Jordan, and Turkey has severely stressed their existing health systems, leaving many Syrian refugees with constrained access to essential health services. For those remaining in Syria, the public health situation is catastrophic. The breakdown of Syria’s public health infrastructure has created an environment ripe for both vaccine-preventable illness, such as measles and polio(7, 8), and communicable disease, most concerningly, COVID-19. There is insufficient availability of testing to know the true extent of the COVID-19 outbreak but there is believed to be widespread community transmission both within government-controlled parts of Syria, non-government controlled sectors in NE and NW Syria and in the crowded informal settlements of Syrian refugees around the region.(9) The degree of food and economic insecurity displaced Syrians are experiencing severely limit any ability to control the spread of COVID-19 through isolation.(9)

Attacks on healthcare workers and facilities have critically impaired health services in Syria.(10, 11) Despite three United Nations resolutions protecting civilians and health care personnel, according to Physicians for Human Rights (PHR) through November 2019, at least 930 health care personnel have been killed in 599 attacks health care facilities since the start of the conflict in March 2011.(12) The dangers facing medical personnel have resulted in the withdrawal of many international humanitarian medical missions from Syria,(13) further worsening Syrians’ ability to access health services.

Trauma care in regions outside the control of the Syrian government and the self-proclaimed Islamic State (non-Government, non-IS; NGNI) was provided by a loose network of established and ad hoc hospitals. While some formal hospitals remained standing, many operated in converted school basements, parking garages, or housing blocks where emergency care was provided with limited inpatient capacity.(14) Injured patients were discharged quickly to convalesce elsewhere due to a lack of inpatient beds and fear of aerial bombardment of NGNI hospitals. While many NGNI hospitals operated under severe human and material resource constraints (e.g., no specialist surgeons, no anesthesiologists, or laypersons functioning as nursing staff), others retained capacity for expanded services including dialysis, intensive care, and diagnostic testing. These larger facilities acted as referral centers for complicated patients and those requiring prolonged inpatient treatment. Ambulances and other ad hoc vehicles transporting patients and medical supplies were frequently destroyed, with drivers detained or killed.(15) Safety of referral and supply routes remained a challenge to clinical operations.

NGNI Syrian hospitals operated independently. Many derived at least partial support from global or regional non-governmental organizations (NGOs) in the form of medications, medical supplies and support for salaries. Coordination between facilities, for resource distribution and referral of patients, was facilitated by local governing councils and a “hospitals committee” associated with the Union of Medical Care and Relief Organizations (UOSSM – French acronym).

Despite coverage in lay press, there remains a paucity of published data on the epidemiology, types and mechanisms of injuries, and associated mortality of trauma patients in the Syrian war. Such analysis is critical to understanding the toll of the war on public health in Syria, to understanding the future needs of the health system at the time of rebuilding, and may have implications for understanding the health impacts of other modern conflicts. This study presents a descriptive analysis of 193,618 trauma patient presentations to 95 hospitals in NGNI-controlled regions of Syria from July 2013 – July 2015. This represents the largest study to date of patient presentations in the Syrian war and provides insights into the civilian impact of the conflict.

## Methods

A retrospective, quantitative secondary analysis was conducted on an administrative dataset of 193,618 trauma patient presentations to 95 NGNI hospitals inside war-affected parts of Syria from July 2013 to July 2015 (Figure 1). The dataset was initially established as an administrative dataset to quantify services and to rationalize material and salary support to facilities providing trauma care in NGNI controlled parts of Syria. Every known such facility was approached for inclusion but not all facilities agreed to share data from the outset in July 2013. Since not all facilities reported every month, an attempt was made to identify which facilities reported in each month and to list causes for non-reporting (Figure 2).

**Figure 1:**
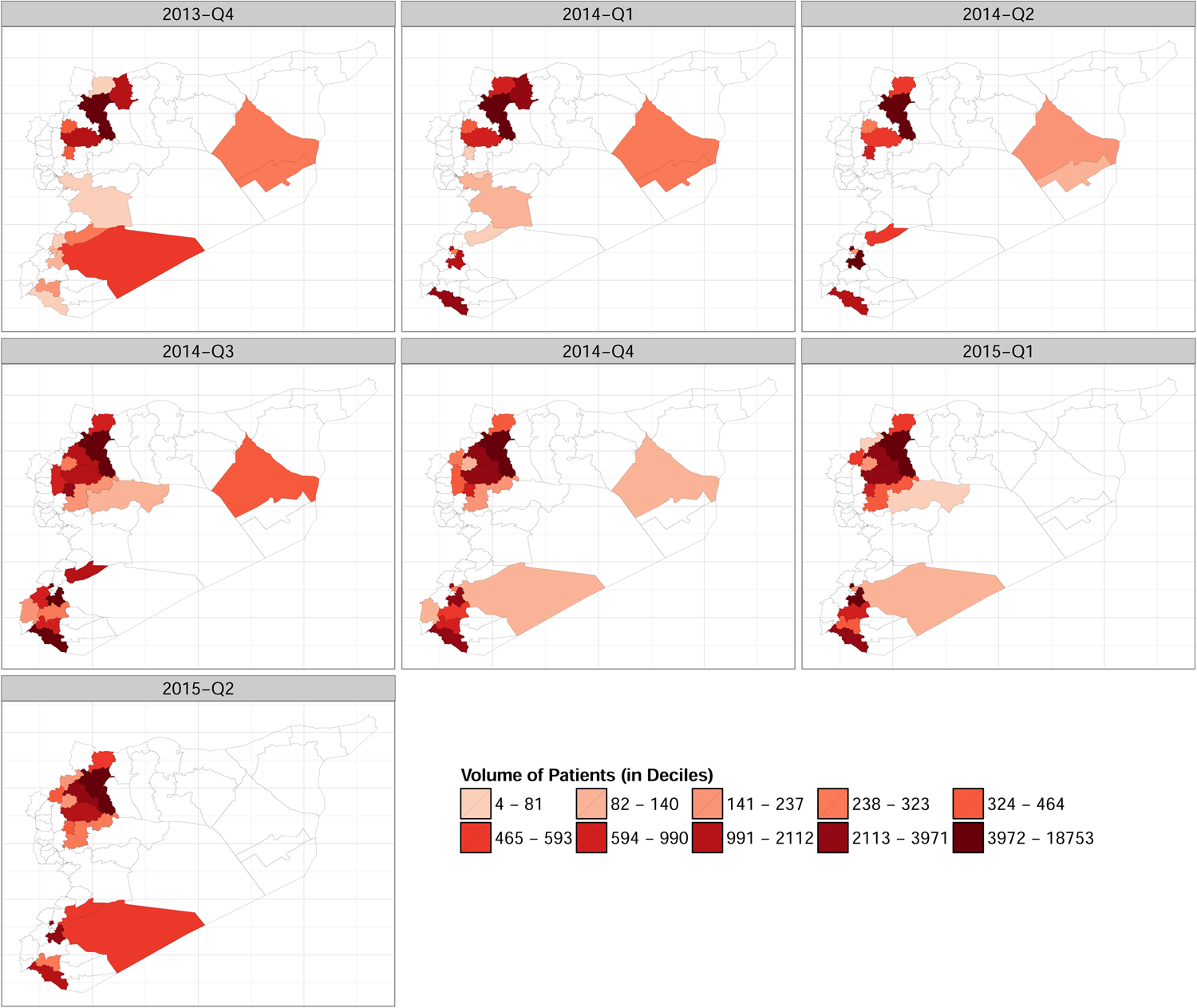
Heat map of war-related trauma patient volume (by quarter)

**Figure 2:**
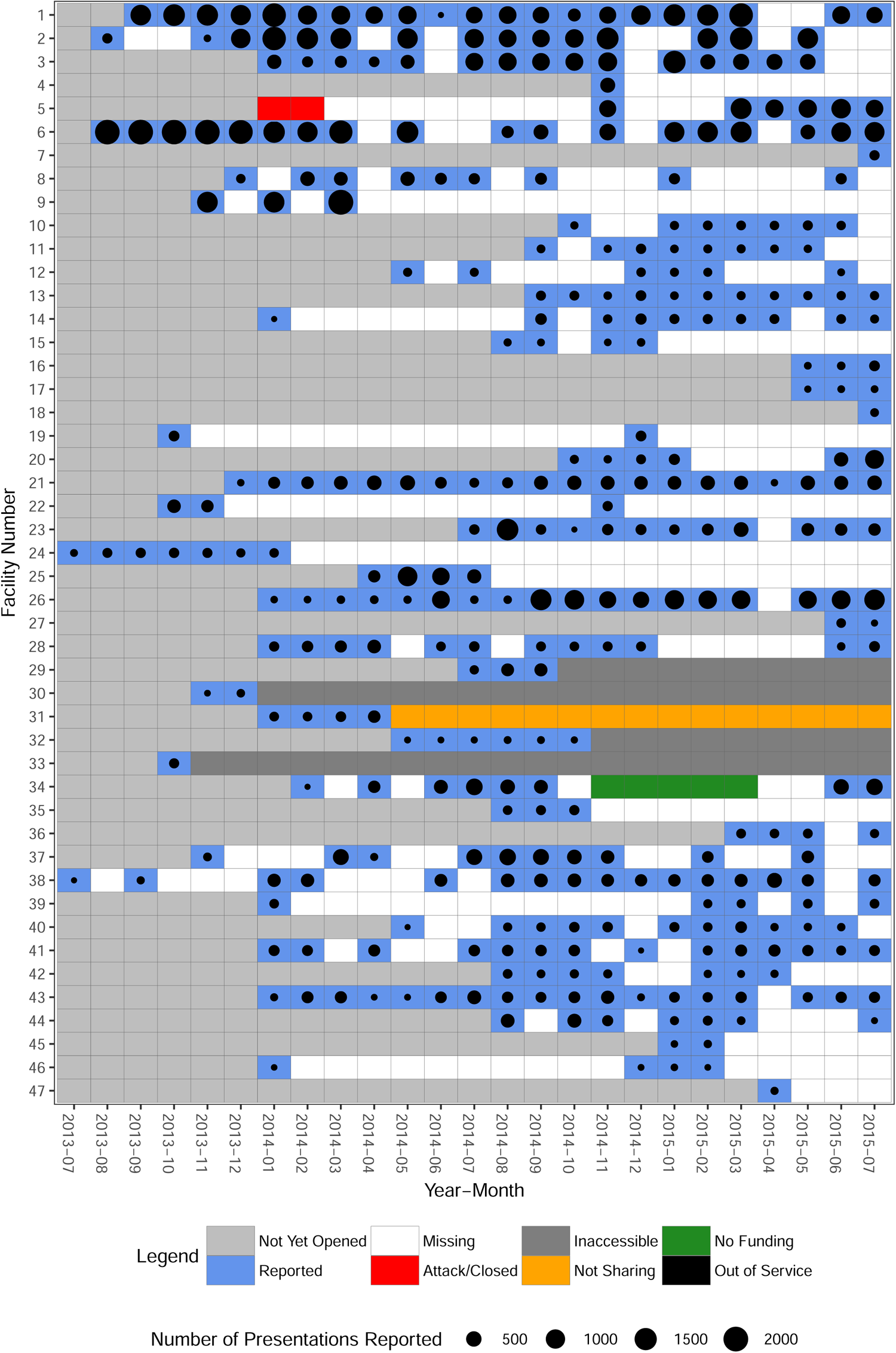
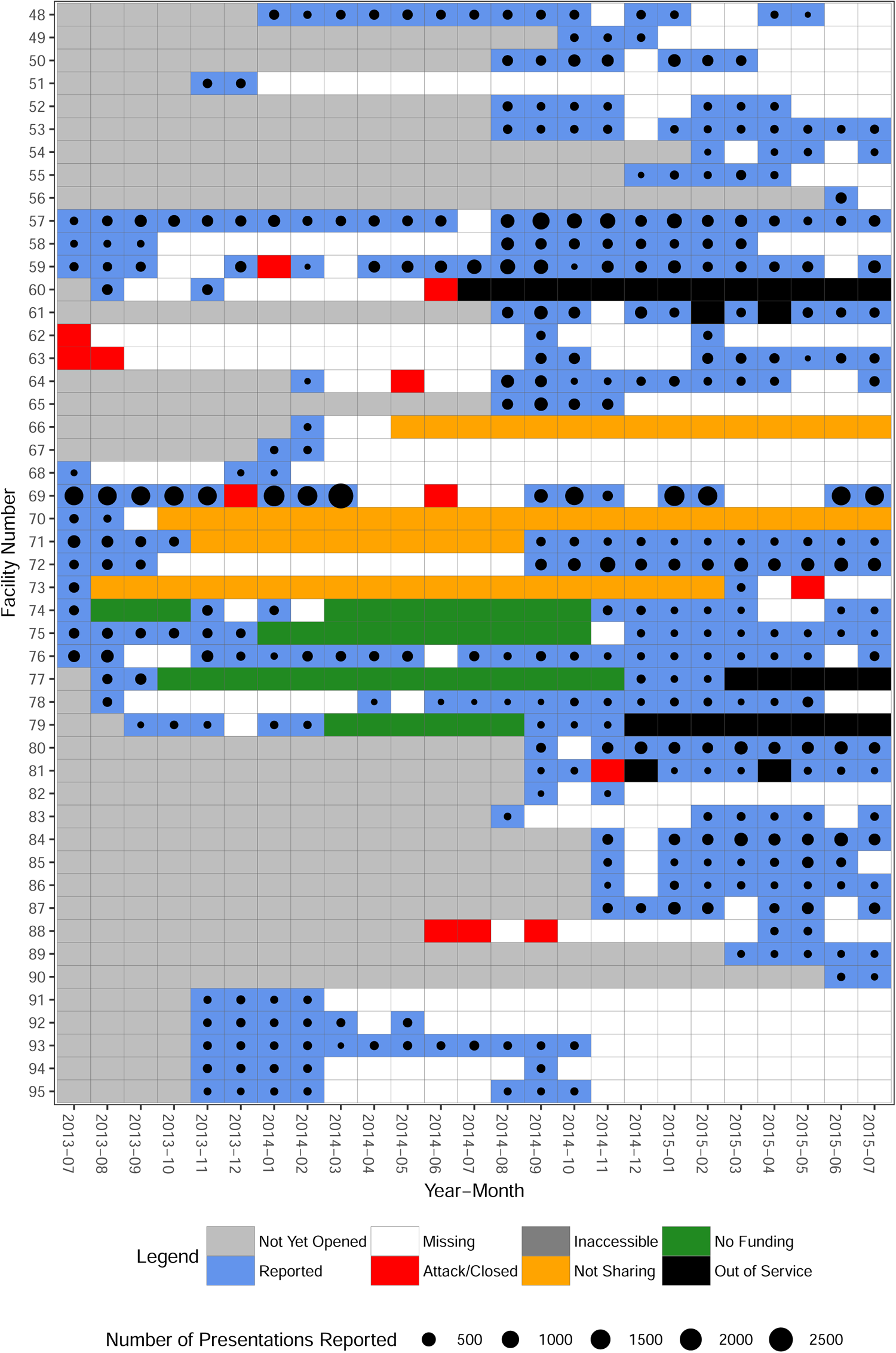
Facility reporting by month and reason for non-report.

### Variables

Variables recorded in the underlying administrative dataset and their parameters were established by the UOSSM hospital committee. They included patient demographics; injury type, location, and mechanism; procedures performed; and patient disposition (Table 1). Specific codes were used for each hospital, procedure type, and injury mechanism.

**Table 1:** Characteristics of Syria War trauma patients (July 1, 2013 to July 31, 2015)

|  | Female |  | Male |  | Total |  |
| --- | --- | --- | --- | --- | --- | --- |
| Total Patient Volume (n, %) | 39393 | 20.4% | 154225 | 79.7% | 193618 | 100% |
| Age Group, years (n, percent) |  |  |  |  |  |  |
| 00-02 | 2587 | 6.6% | 5670 | 3.7% | 8257 | 4.3% |
| 03-12 | 7123 | 18.1% | 17076 | 11.1% | 24199 | 12.5% |
| 13-18 | 4222 | 10.7% | 18260 | 11.8% | 22482 | 11.6% |
| 19-30 | 8467 | 21.5% | 51504 | 33.4% | 59971 | 31.0% |
| 31-40 | 4515 | 11.5% | 17537 | 11.4% | 22052 | 11.4% |
| 41-50 | 3175 | 8.1% | 8642 | 5.6% | 11817 | 6.1% |
| 51-60 | 2125 | 5.4% | 4588 | 3.0% | 6713 | 3.5% |
| >60 | 1493 | 3.8% | 3253 | 2.1% | 4746 | 2.5% |
| Unknown/Missing Age | 5686 | 14.4% | 27695 | 18.0% | 33381 | 17.2% |
| Injury Mechanism (n, %) |  |  |  |  |  |  |
| Blast | 509 | 1.3% | 3334 | 2.2% | 3843 | 2.0% |
| Blunt/Crush | 20204 | 51.3% | 51273 | 33.3% | 71477 | 36.9% |
| Burn | 1640 | 4.2% | 4216 | 2.7% | 5856 | 3.0% |
| Gunshot | 2513 | 6.4% | 19827 | 12.9% | 22340 | 11.5% |
| Shrapnel | 12278 | 31.2% | 69668 | 45.2% | 81946 | 42.3% |
| Stab Wound | 1290 | 3.3% | 4422 | 2.9% | 5712 | 3.0% |
| Wound | 672 | 1.7% | 2231 | 1.5% | 2903 | 1.5% |
| Trauma Other | 529 | 1.3% | 1724 | 1.1% | 2253 | 1.2% |
| Surgical (Non-Trauma) | 15 | 0.04% | 32 | 0.02% | 47 | 0.02% |
| Surgical Procedures |  |  |  |  |  |  |
| Total (n) | 41265 |  | 168505 |  | 209770 |  |
| Per Patient, All (mean, SDev) | 1.05 | 0.39 | 1.09 | 0.43 | 1.08 | 0.43 |
| Per Patient, Admitted (mean, SDev) | 1.13 | 0.66 | 1.25 | 0.67 | 1.23 | 0.67 |
| ED Disposition (n, %) |  |  |  |  |  |  |
| Discharge from ED | 28308 | 72.4% | 103792 | 67.3% | 132300 | 68.3% |
| Admit | 10311 | 26.2% | 47314 | 30.7% | 57625 | 29.8% |
| Admit ICU | 246 | 0.6% | 1516 | 1.0% | 1762 | 0.9% |
| Death | 328 | 0.8% | 1603 | 1.0% | 1931 | 1.0% |
| Length of Stay – median (days, SDev) | 4.52 |  | 3.59 |  | 3.80 |  |
| Final Disposition – admitted (n,%) |  |  |  |  |  |  |
| Discharge from Hospital | 9136 | 86.5% | 41799 | 85.6% | 50935 | 85.8% |
| In Hospital | 27 | 0.3% | 220 | 0.5% | 247 | 0.4% |
| Transfer to Other Facility | 659 | 6.2% | 4099 | 8.4% | 4758 | 8.0% |
| Death | 601 | 5.7% | 2093 | 4.3% | 2694 | 4.5% |
| Unknown | 134 | 1.3% | 617 | 1.3% | 751 | 1.3% |

### Reporting

NGNI hospitals sent monthly data along with characteristics of facilities (Table 2) to the UOSSM hospital committee that was used to assess the overall volume of major cases across facilities. Data were transcribed from handwritten hospital logs into encrypted Microsoft Excel spreadsheets (Microsoft Corporation (2013), Redmond, Washington) and sent for inclusion in a central database running Microsoft SQL Server software (Microsoft Corporation (2012) Redmond, Washington). In addition, monthly facility visits were made by a member of the hospital committee data collection team (starting March 2014) to review offline records and to ensure no major discrepancies from what was reported electronically.

**Table 2:** Characteristics of Syrian Trauma Hospitals (July 1, 2013 to July 31, 2015)

|  |  |  |
| --- | --- | --- |
| Total number of facilities reporting (n) |  | 95 |
| Patient Volume Total, monthly per facility (mean) |  | 267.43 |
| Patient admissions, monthly per facility (mean monthly) |  | 83.64 |
| Geographic distribution of presentations<br>(mean monthly) | Per 100K* | Total |
| Aleppo | 80.4 | 4090.6 |
| Damascus | 5.3 | 116.6 |
| Damascus (Rural) | 836.9 | 1191.8 |
| Dar'a | 171.5 | 832.1 |
| Deir Ezzor | 11.0 | 153.7 |
| Hama | 30.1 | 406.9 |
| Homs | 1.5 | 35.0 |
| Idleb | 51.4 | 1203.9 |
| Quneitra | 106.2 | 86.0 |
| * Strategic Needs Analysis Project (SNAP) (2015). REGIONAL ANALYSIS Q4 2014 \| 01<br>OCTOBER - 31 DECEMBER 2014. <u>Regional Analysis for Syria</u>. ACAPS. Geneva,<br>ACAPS: 54. |  |  |

### Data preparation

Records were de-identified by the UOSSM system administrator prior to sharing with study investigators. The dataset was cleaned and rare records of non-traumatic presentations (e.g. caesarean section or appendectomy) were removed. The data were subsequently analyzed to characterize the epidemiology of trauma patients, the major causes of presentations and admissions, as well as factors associated with inpatient mortality.

There was variation in the format of age data in the administrative data set including date of birth, age, estimated age, and age group. The cutoffs for age groups did not correspond to international norms but were determined by the NGNI hospitals themselves – infants and toddlers (age 0-2); pre-adolescent children (age 3-12); teenagers (age 13-18); and then roughly to decades of life thereafter with elderly greater than 60 years of age grouped together. In the secondary analysis, a unified age variable was created for analysis and the least granular constituent variable (age group) dictated cutoffs for the combined age variable.

Some facilities, but not all, recorded an initial disposition from the Emergency Department (ED). In order to normalize data across all facilities, an ED disposition was imputed using a pre-specified, rule-based procedure (Appendix A).

### Analysis

Descriptive statistics regarding ages of injured patients, mechanisms of injury, and types of injuries sustained were calculated in R version 3.2.2(16) and data visualization was done using the “ggplot2” package in R.(17)

To predict inpatient mortality based on patient characteristics (such as gender, age group, and injury type), we employed Firth’s biased-reduced logistic regression using the “logistf” package in R.(18) Logistic regression is frequently used to predict binary outcomes and we employed Firth’s bias-reduced logistic regression because death is very rare for patients with certain characteristics in these data. This method is recommended for analyzing rare events as it produces finite parameter estimates and avoids those approaching infinity in absolute value.(19) We performed both univariate regression analysis where odds ratio (OR) for inpatient mortality were reported unadjusted for other characteristics, and multivariate regression analysis where adjusted odds ratio (aOR) for inpatient mortality were reported after adjusting for gender, age group, and injury mechanism-body part combination. Analysis was performed on all records, as well as separately for females and males. In the multivariate regression, the largest age group (19 – 30 years) was chosen as the reference group.

The institutional review board of Yale University approved this study.

## Findings

### Demographics

Over the 25-month study period, 193,618 patient presentations were made to these 95 NGNI hospitals (Table 1, Figure 3). Of these, 154,225 were male (79.7%) and 39,393 female (20.4%). Age information was complete for 160,237 encounters (82.8%). Patients aged 19-30 years (33.4% males, 21.5% females, 31.0% overall, Figure 3) represented the largest single group of patients. Together, children 0-18 years (54,938, 28.4%) and the elderly over 60 years of age (4,745, 2.5%) constitute over one-third of all patients presenting with traumatic injuries (37.2% of patients with known ages).

**Figure 3:**
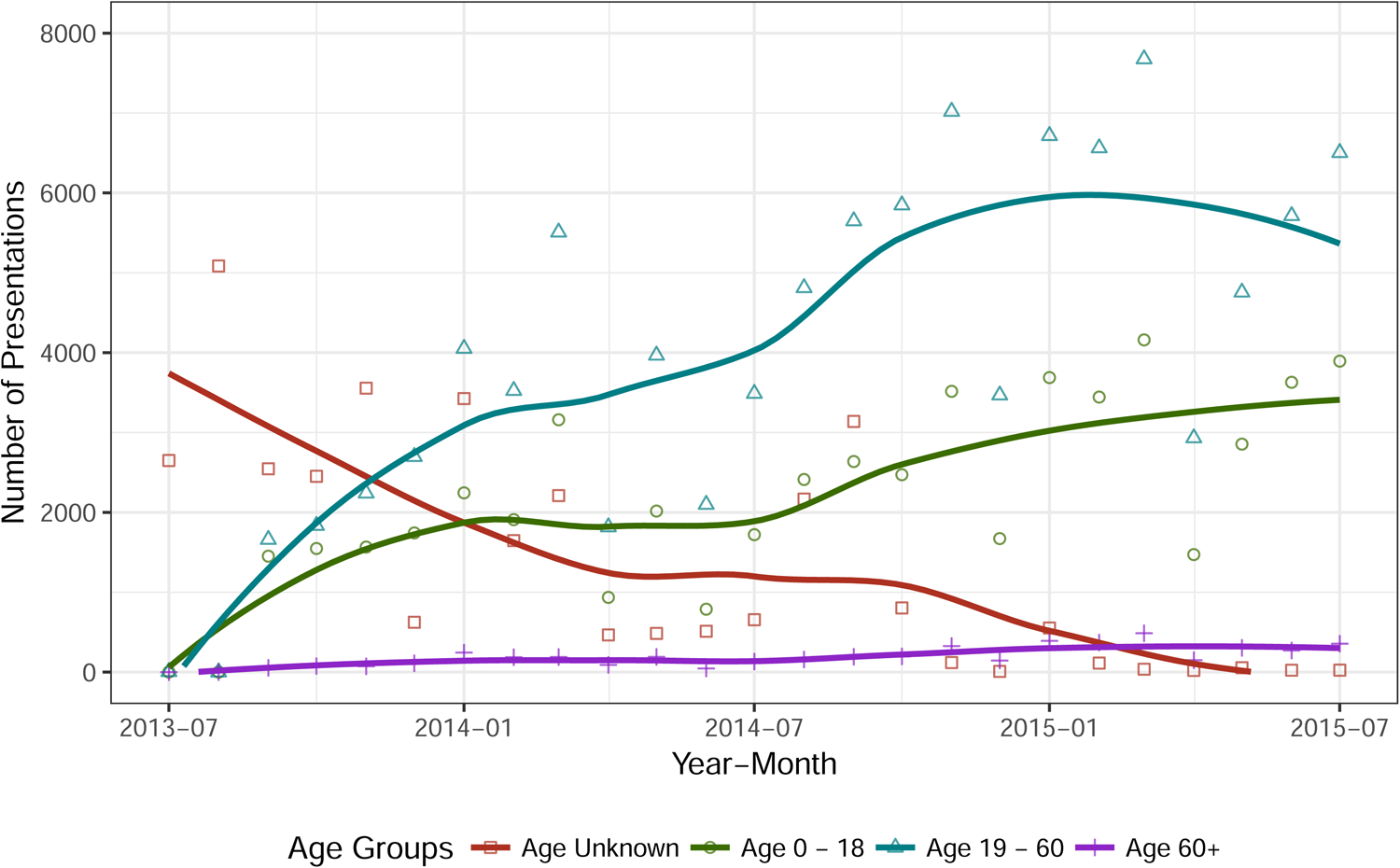
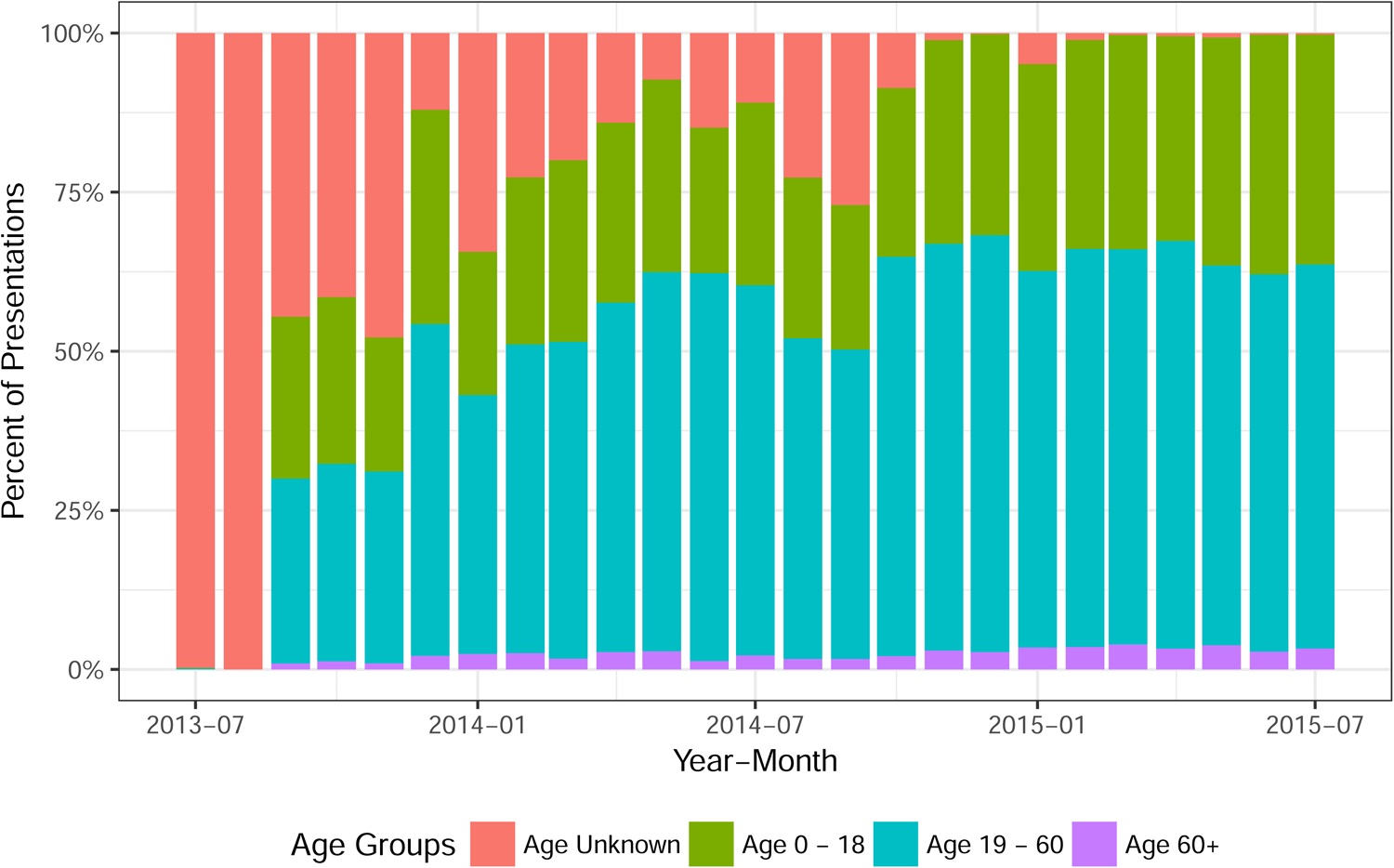
a) Age distribution of war-trauma patients over time; b) as percentage.

### Hospital Reporting

The numbers of patient presentations (Figure 3) and of NGNI hospitals reporting trauma data (Figure 2) varied over time with a continually increasing trend in both. The governorates of Aleppo and Idleb had the highest monthly volume of trauma patient presentations (Table 2, Figure 1) while Rural Damascus and Dar’a governorates were hardest hit in terms of presentations per 100,000 erstwhile population (based on estimate of mid-study population 2014Q4 - Table 2).(20) Besieged areas in Rural Damascus saw an especially high number of patient presentations with 836.9 monthly patient presentations per 100,000 erstwhile population.(20)

### Injury Types

There were 81,946 “shrapnel” injuries (42.3%) (“shrapnel” as recorded in clinical records) and 71,477 blunt/crush injuries (36.9%) which together represented the predominant injury mechanisms overall. Gender differences were noted with blunt/crush injury being the leading mechanism for females (20,204; 51.3%) and shrapnel being the leading mechanism for males (69,668; 45.2%). (Figure 4)

**Figure 4:**
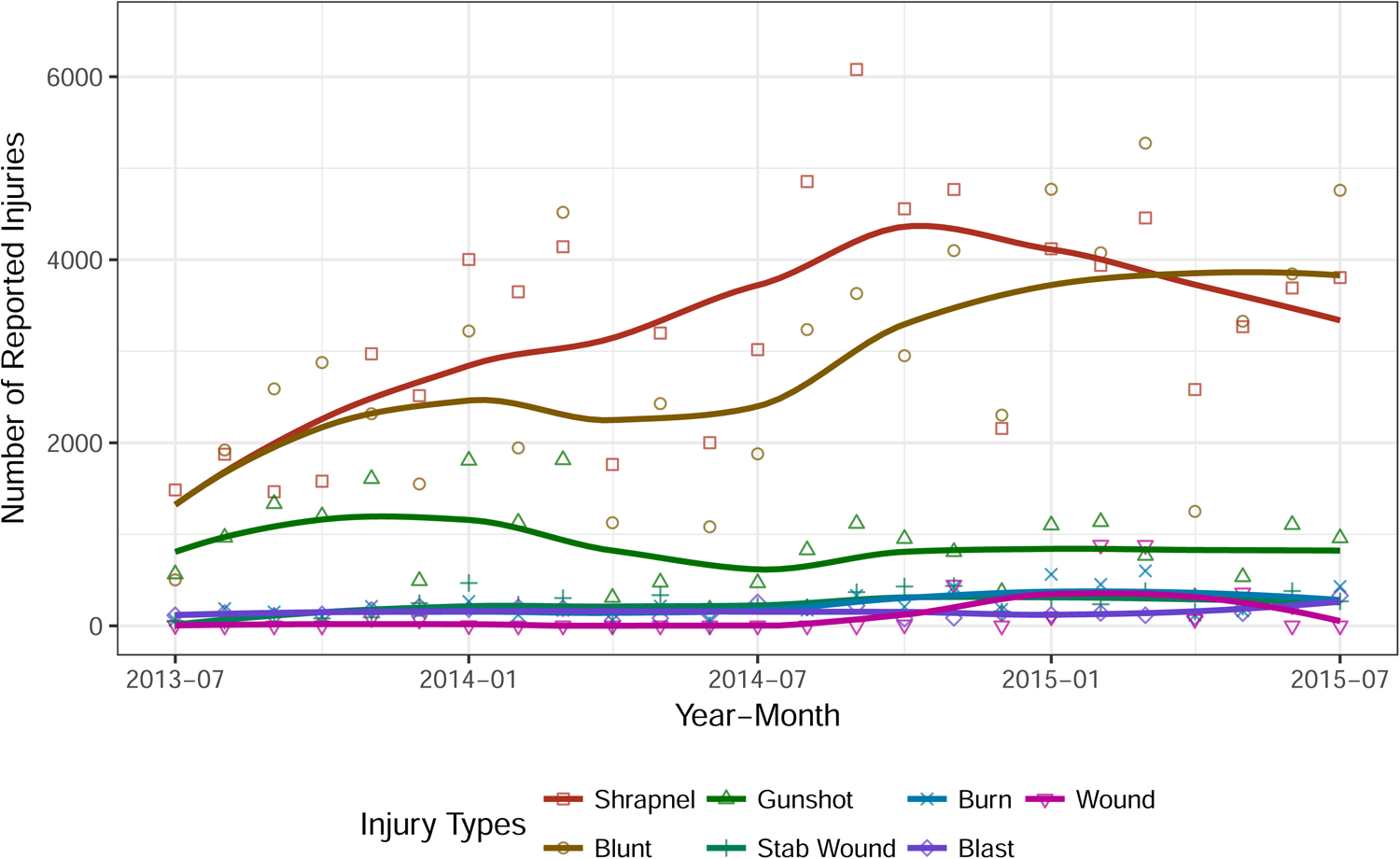
Injury mechanisms over time for Syrian war trauma patients.

### Procedures

There were 209,770 procedures documented resulting in 1.08 (SD 0.43) procedures per patient visit. Patients discharged from the ED primarily underwent minor surgical or bedside procedures (e.g., reduction and immobilization of fractures, laceration repair, or abscess drainage). Admitted patients primarily underwent procedures that require general anesthesia and the operating theater.

### Disposition

A total of 59,387 (30.7%) admissions were recorded (57,625 wards, 29.8%; 1,762 ICU, 0.9%) for an average length of stay of 3.8 days. Many facilities lacked formal critical care capacity. Critical care or ICU care was noted only when a critical care intervention was noted (e.g., mechanical ventilation) or when a critical care admission code was recorded.

### Mortality

Of admitted patients, 50,935 (85.8%) survived to discharge and 4,758 (8.0%) were transferred to another facility for continued care. There were 4,625 facility-based deaths recorded (1,931 in Emergency Department and 2,694 inpatients). Overall mortality rate was 2.4% (4.5% for admitted patients).

Factors most associated with mortality included: extremes of age (age less than 2, aOR 2.92; age greater than 60, aOR 2.48) and injury mechanism-body part combinations including: blast-head (aOR 13.43); blast-spine (aOR 11.31); gunshot-head (aOR 10.07); “shrapnel”-head (aOR 6.34); and gunshot-chest (aOR 6.03) (Table 3). Similar patterns were seen for admitted patients with neurotrauma and penetrating injuries of head and chest accounting for highest associations with mortality. Blunt/crush-chest had a more significant association with mortality in admitted patients than in the overall population (Appendix B).

**Table 3:**
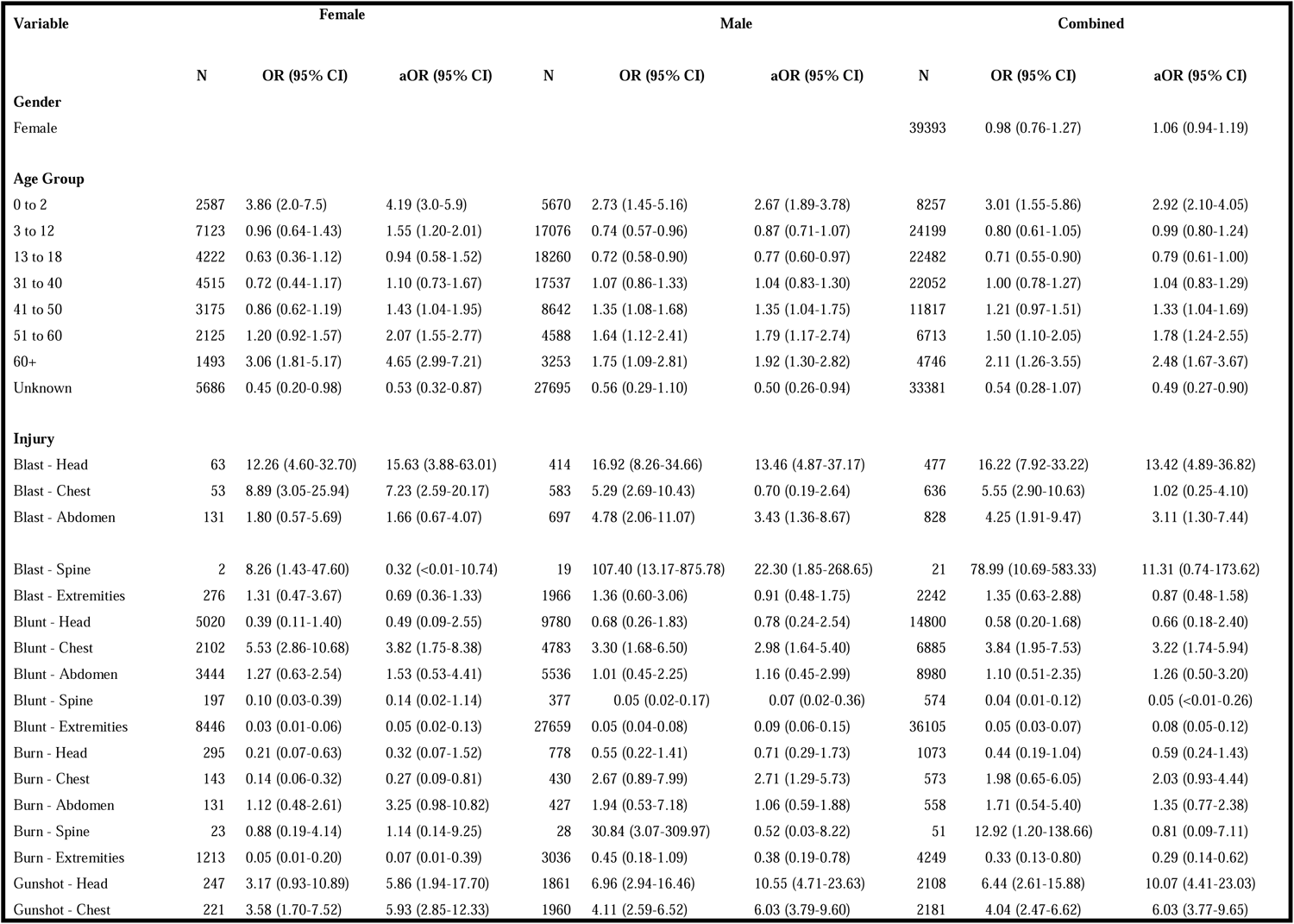
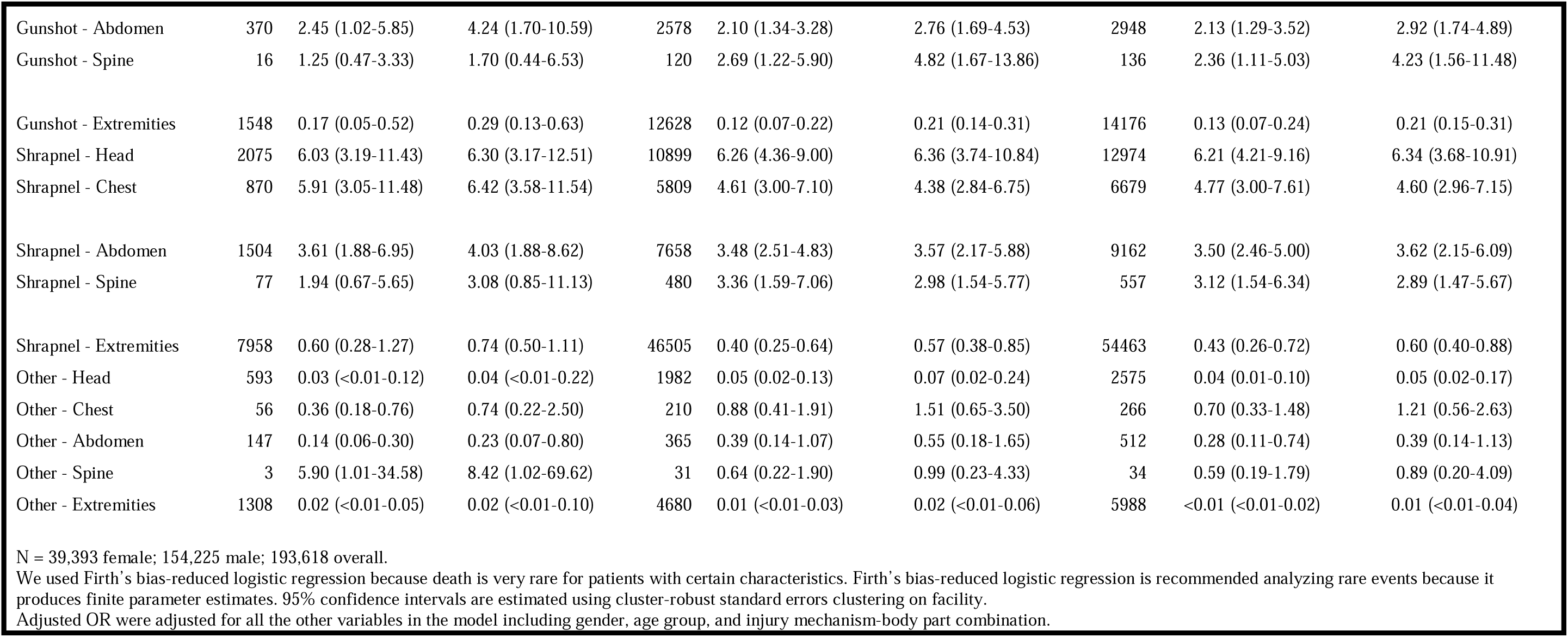
Predicting Death Among All Presentations Using Firth Logistic Regression.

## Interpretation

This study of 193,618 trauma presentations from 95 NGNI Syrian hospitals providing trauma care in regions outside Syrian government control represents the largest analyses of trauma presentations in the Syrian conflict to date.(21–31) Dramatic increases in mortality from the Syrian war have resulted in a drop of life expectancy to a low of 59.4 years in 2014 compared to expected 68 years.(32) Civilians were heavily impacted by war-related trauma in Syria. A 2018 article by Guha-Sapir et al demonstrated that 70.6% of war deaths documented between 2011-2016 were civilians.(27) The findings of our study of inpatient mortality further confirm the civilian impact of the conflict.

Using the UN Convention on the Rights of the Child definition of a child being, “a person below the age of 18…” our study reveals that children, women and elders make up a large and increasing proportion of the war-injured presenting to NGNI Syrian hospitals rising to a peak of 51% at the end of data collection (Figure 5). While the total number of child combatants in the Syrian conflict is unknown, in 2012 the Violations Documentation Center documented 194 “non-civilian” child deaths since 2011.(33) In 2014, Human Rights Watch documented male child combatants as young as 16 year of age and males as young as 14 years old serving in support roles with armed groups. It was estimated that children represented 2% of child deaths in the conflict.(34) There are much fewer cases of female child combatants in the Syrian conflict. While teenage females were documented fighting with the YPJ (Kurdish forces female units), the Kurdish forces signed a “deed of commitment” with the non-governmental organization Geneva Call to ban the use of all child soldiers in 2014.(35) While reports exist of child soldiers on all sides in the Syrian conflict, using the most conservative assumption that *every male* 13-60 years of age and that *every single male* of unknown age was a combatant – civilians still would constitute a large and increasing burden of trauma patients with a peak under this extremely conservative model of 39.6% of cases (Figure 5) suggesting a very high burden of civilian trauma in the Syrian war.

**Figure 5:**
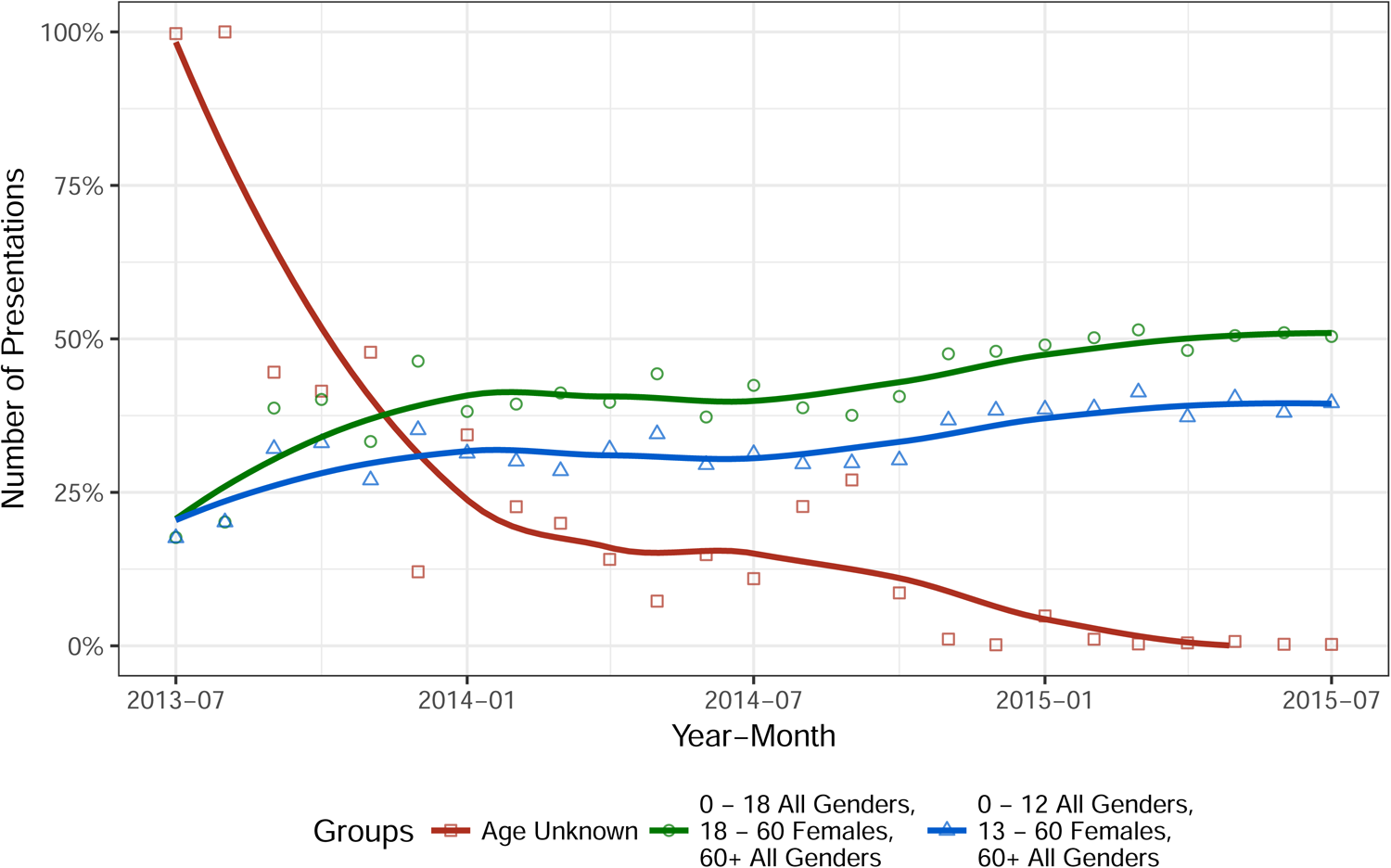
Presumed civilian war trauma injuries over time.

Males aged 19-30 represented the largest single group of patients in this study. While some in this age group may have been injured as combatants, this group may also be over-represented in the population remaining within Syria due to disproportionate refugee displacement of children, women, and elders outside the country.(36)

Gunshot wounds remained a major cause of presentations (third overall). The number and proportion of injuries caused by blunt/crush injuries as well as penetrating injuries from flying metal fragments increased significantly over time, including among children and the elderly, while numbers of injuries from other mechanisms remained constant(Figure 4). This pattern is consistent with the transition to predominance of aerial bombardment with missiles and barrel bombs resulting in building collapse in the later years of the conflict. The association of “blast” and “shrapnel” injuries with mortality ranged from less than 1 to greater than 13 – highest for neurotrauma (head and spinal injuries). This is consistent with the limited capacity of NGNI hospitals to provide specialty care for these devastating injuries.(37)

The median length of stay for admitted patients was 3.8 days. This very brief median duration of hospitalization may reflect the limited capacity of NGNI hospitals in Syria for inpatient care.(37) Anecdotally, data collectors relayed concerns of hospital staff that facilities were targeted by armed forcese and that patients were routinely discharged to convalesce from their injuries elsewhere rather than maintain large numbers of patients in one place.

The patterns of injury and mortality we report comport with results of previous studies of mortality in modern wars. While in most studies males of fighting age represent the bulk of the killed and injured,(25, 27–29, 38–40) Speigel and Salama demonstrated increased mortality amongst males greater than 50 years of age in their epidemiological study of war mortality in the Kosovo war of the early 1990s(38) and a review of civilian injuries in modern conflicts by Aboutanos and Baker revealed a peak of civilian casualties in the elderly population.(39) More recent studies of the wars in Iraq, Afghanistan and Syria also document the transition from gunshot wounds predominating at the start of the conflict to the higher prevalence of injuries from explosive ordinance.(27, 28, 39, 40)

## Limitations

Many of the limitations of this study are a function of the limitations of the underlying administrative dataset from which it is derived. The original administrative dataset’s purpose was to quantify the added burden of “war trauma patients” on facilities for resource allocation. As such, data collected emphasized a) more significant injuries (lesser injuries and non-operative injuries frequently not recorded when occurring concurrently with major ones); and b) bias towards intentional and war-related trauma (unintentional injuries and routine household trauma were under-reported). One must assume that routine falls and motor vehicle collisions, for example, continued to occur despite ongoing hostilities yet they are largely absent from this administrative dataset. With this in mind, the results presented here likely represent an under-estimate of the true number of trauma presentations in the Syrian war. Future modifications of this data system should capture at least a minimal set of data on all patients to more accurately reflect the clinical load and resource utilization associated with trauma care.

In addition, injury mechanisms reported here are taken directly from the administrative record as recorded. We report mechanisms as they were recorded by the treating physicians. However, the use of outdated terminology such as “shrapnel” (as opposed to “secondary injury due to flying objects”) and “blast” (when it cannot be determined if they are referring only to injuries due to overpressure) limits comparison to other studies where injuries are more precisely characterized.

Additionally, there was no ability to uniquely identify patients in a verifiable sense between NGNI hospitals. As such, there was no ability to tie complications (such as infection or bleeding) to patients’ previous initial presentations. Similarly, the inability to follow up patients after discharge meant that out-of-hospital deaths occurring after discharge are absent from these data and likely contribute to the very low documented mortality in this report of war trauma. Future efforts should incorporate methods to dynamically assign chart numbers in a decentralized but systematized fashion that can be consistently applied throughout hospitals in the network with use of assigned numbers on return visits to better capture medium- and long-term outcomes.

NGNI Syrian hospitals in this study operated under immense security risks and resource constraints. Many facilities included in this analysis were damaged or destroyed (some more than once) during the course of the study period. In addition, since coming under the control of the self-proclaimed Islamic State (IS), hospitals in the east of the country ceased to contribute to the administrative database. In some cases, it is not known whether some facilities were destroyed or simply stopped sharing data. Facilities were only noted as closed if they were confirmed as destroyed or closed. Otherwise, they were recorded as “missing” data.

Further, data collection in 2013 and early 2014 was largely passive and voluntary. In spring 2014, the hospital committee instituted monthly site visits to verify reported data and to report back concerns from the facilities to the hospitals committee. This active surveillance resulted in an immediate uptick in the number of patient presentations reported which continued throughout the remaining study period. The number of NGNI hospitals reporting monthly in the underlying administrative dataset varied over time. We have attempted to clearly illustrate when each facility reported and, when possible, causes for missing data (Figure 2).

There may be NGNI hospitals that were operating in this period that are completely absent from this analysis. Previous surveys of NGNI hospitals in the region revealed 93 hospitals with operating theaters (HeFRA-2)(41), 59 hospitals and 86 “secondary health facilities” (HeRAMS)(42), a spring 2015 nationwide survey of trauma hospitals reported 86 hospitals with the ability to “perform emergency surgery”(37), and most recently a March 2017 hospital survey that revealed 107 facilities providing operative care (26 formal hospitals, 43 operating in schools or public buildings, 34 field hospitals, and 4 underground/caves)(14). As such, while there is no exhaustive list of operative facilities in these regions and the destruction and reconstitution of facilities occurs regularly in this conflict – the 95 NGNI hospitals reporting data here represented the majority of operative trauma care facilities in these areas.

Isolation and the constant threat of bombardment made data collection, transmission and verification challenging. There were no functioning phone or data networks in most areas of interest during the reporting period with most communication carried out by two-way satellite connections. Reporting delays in the underlying administrative dataset were frequent and verification only took place when security conditions permitted safe passage.

These data reflect only *inpatient* mortality. Patients who died pre-hospital, shortly after arrival, or after discharge are absent from these data. There may be a survivor bias in these results. Mortality causes and overall demographic distribution may have been skewed with those who survived to hospital presentation having less severe injuries or different demographic profile than overall war-injured. Critically important, these inpatient data cannot provide estimates of the population level impact of injuries in the Syrian war as they by definition exclude those that could not reach health services. These factors would result in an underestimate of both the number and acuity of patient presentations.

The overall in-hospital mortality rate was 4.5%, a number more reflective of a civilian trauma center in peace-time(43) than a network of under-resourced hospitals during war. This likely represents under-recording of those who died a) immediatelyt at the scene; b) during prehospital transport; or c) shortly after arrival. According to those authors with experience providing trauma care inside Syria during this war, when large numbers of severely injured patients arrived simultaneously those with minor injuries, those who were peri-morbid, and those who were dead on arrival frequently had no clinical chart generated. While it is understandable that providers over-stretched with patient care were reluctant to add to their administrative load for patients deemed unsalvageable, care must be taken in future data collection to register such patients and to capture a minimum dataset based on established injury surveillance guidelines emphasizing demographic factors, vital status and details of major injuries to more accurately reflect the mortality rates associated with these trauma presentations.(44)

Recorded ICU admissions in this dataset represent an underestimate of the need for critical care. ICU admission was only noted when an ICU admission code was used and/or a critical care intervention was noted (e.g., mechanical ventilation). As such, NGNI hospitals that provided critical care but did not have a formal ICU are not reflected in this dataset unless they explicitly used a procedure code for a critical care intervention in the administrative dataset. Future data collection should incorporate simple measures to highlight critical patients prospectively and to ensure that similar coding practices are utilized across the network.

Date of discharge was frequently missing. Few NGNI hospitals had significant inpatient capacity and most patients were released within one day, even after surgical intervention under anesthesia, for reasons explained above.

Routinely, records with missing discharge dates were assigned length of stay of one day in the base administrative dataset. Such modifications were made prior to provision of the underlying dataset to the study investigators. As such, there exist an unknown number of records for which the length of stay may be recorded as shorter than the patient’s actual length of hospitalization. Future improvements should employ data validation tools to either force recording the discharge date or, for example, to use the date of data entry obtained from the device clock for records entered electronically.

Our results are further limited by the lack of a prospectively applied injury severity score. Future data collection should incorporate an estimate of injury severity to better describe the burden of injury represented in the data, guide quality of care efforts, and identify hospitals with best practices and those in need of additional support.

## Conclusion

Civilians were heavily impacted by war-related trauma in Syria. Children, women and the elderly together make up the major portion of Syrian war trauma patients comprising more than half of all patients in the final months of this study. It is important to note that this rising trend in the data presented here also precede the heavy aerial bombardment of Aleppo and other cities in Northern Syria Syrian Government and allied forces in the summer and autumn of 2015. Over the period captured in this study, the shift in injury patterns from predominance of gunshot wounds to blunt/crush injuries and penetrating injuries from explosive fragments mirrored the widely reported shift to predominance of aerial bombardment over the course of the war. Future work should emphasize standardization of data variables and capturing a minimum dataset on all trauma patients to improve estimates of mortality and its causes in the Syrian civil war. Simple modifications to the hospital data collection schemes (e.g., inclusion of a simple discharge summary) can enhance the ability to capture hospital complications that may not be apparent or recorded at the time of the initial trauma documentation.

This paper presents the largest dataset of trauma presentations in the Syrian war – several magnitudes greater than other facility-based data presented in the literature(21–31) – and one that realistically reflects the limitations of data acquisition and patient tracking in war. Further, this study serves to highlight areas where data quality improvements can inform future data collection efforts in Syria and other conflict-affected settings.

## Data Availability

The data for this study is available upon request from the authors.

## Acknowledgements

The authors acknowledge the health workers who, at great personal risk, provide life-saving care for the sick and injured inside Syria and the data collection team who similarly faced threats to their safety collecting the data that informed this study. In addition, the authors would like to recognize the 95 hospital directors who provided access to their data and the partner organizations including the Union of Medical Care and Relief Organizations (UOSSM), Syrian American Medical Society (SAMS), Physicians Across Continents (PAC), Syrian Expatriate Medical Association (SEMA), Sham Humanitarian Fund, and all the other members of the hospital committee and the independent NGNI hospitals inside Syria whose cooperation enabled this report. Finally, the authors would like to express our thanks to Dr. Monzer Yazji for helping to facilitate this research collaboration.

## Declaration of Interests

The authors would like to state that they have no financial conflicts to disclose. Three of the authors have worked inside Syria as physicians providing direct medical care (MH, HA, AA).

| Author Name | Affiliation | Role/Contribution |
| --- | --- | --- |
| Hani Mowafi* | Yale University | Drafting Manuscript,<br>Study conception,<br>Study design,<br>Data Management,<br>Planned Analyses,<br>Final Review |
| Mahmoud Hariri* | UOSSM | Study design,<br>Data Management,<br>Final Review |
| Baobao Zhang | Yale University | Statistical Analyses,<br>Data visualization,<br>Methods<br>Final review |
| Houssam Alnahhass | Istanbul University<br>UOSSM | Data Management,<br>Final Review |
| Basil Bakri | UOSSM | Data Management,<br>Methods,<br>Final Review |
| Adam Eldahan | Columbia University | Background,<br>Data Management<br>Literature research<br>Final Review |
| Moustafa Moustafa | University of<br>Virginia | Background,<br>Literature research<br>Final Review |
| Maher Saqqur | University Alberta | Analysis consultation<br>Final Review |
| Anas Alkassem | UOSSM | Senior Author,<br>Conception of study,<br>Data Management,<br>Final Review |

\* Co-First Authors

## References

1. Syrian Observatory for Human Rights. Syrian Observatory for Human Rights London: Syrian Observatory for Human Rights; 2019 [Available from: http://www.syriahr.com/en/?p=120851.

2. Ohlheiser A. The U.N. Has Stopped Counting the Deaths In Syria. The Atlantic. 2014 07 January 2014.

3. Lancet Editorial. The war on Syrian civilians. Lancet. 2014;383(9915):383.

4. Alexandra Bilak MC, Guillaume Charron, et al. Global Overview 2015: People internally displaced by conflict and violence. Geneva: Norweigian Refugee Council Internal Displacement Monitoring Centre (IDMC); 2015.

5. UNHCR. Syria Emergency Geneva: UNHCR; 2020 [updated 15 March 2021. Available from: https://www.unhcr.org/en-us/syria-emergency.html.

6. UNHCR. Syria Regional Refugee Response Geneva: UNHCR; 2020 [updated 31 March 2021. Available from: https://data2.unhcr.org/en/situations/syria.

7. Sharara SL, Kanj SS. War and infectious diseases: challenges of the Syrian civil war. PLoS pathogens. 2014;10(10):e1004438.

8. Friedrich M. Vaccination Effort Targets Middle East After Polio Outbreak in Syria. JAMA. 2013;310(23):2497-.

9. iMMAP. COVID-19 SITUATION ANALYSIS CRISIS TYPE: EPIDEMIC-Syria February 2021. 2021 February 2021.

10. Anatomy of a Crisis: A Map of Attacks on Health Care in Syria [Internet]. Physicians for Human Rights. 2015 [cited Aug 4, 2015]. Available from: https://s3.amazonaws.com/PHR_syria_map/web/index.html.

11. Hampton T. Health Care Under Attack in Syrian Conflict. JAMA. 2013;310(5):465–6.

12. PHR: Physicians for Human Rights. Anatomy of a Crisis: A Map of Attacks on Health Care in Syria New York: Physicians for Human Rights; 2021 [updated 31 March 2021. Available from: https://s3.amazonaws.com/PHR_syria_map/web/index.html.

13. Arie S. MSF charity closes medical facilities in parts of Syria after staff were kidnapped. BMJ. 2014;348.

14. Union of Medical Care and Relief Organizations (UOSSM). Syrian Hospitals Surveillance Study. Geneva: Union of Medical Care and Relief Organizations (UOSSM),; 2017 March 2017.

15. United Nations General Assembly Human Rights Council. Report of the Independent International Commision of Inquiry on the Syrian Arab Republic. United Nations General Assembly Human Rights Council; 2015 5 February 2015.

16. R Core Team. R: A language and environment for statistical computing. In: Computing RFfS, editor. Vienna, Austria 2013.

17. Wickham H. ggplot2: Elegant Graphics for Data Analysis. New York: Springer-Verlag; 2016.

18. Georg Heinze MP, Daniela Dunkler, Harry Southworth. logistf: Firth’s Bias-Reduced Logistic Regression. Vienna 2018.

19. Heinze G, Schemper M. A solution to the problem of separation in logistic regression. Statistics in Medicine. 2002;21(16):2409–19.

20. Strategic Needs Analysis Project (SNAP). Regional Analysis Q4 2014 | 01 October - 31 December 2014. Geneva: ACAPS; 2015 Feb 2015.

21. Hakimoglu S, Karcıoglu M, Tuzcu K, Davarcı I, Koyuncu O, Dikey İ, et al. Assessment of the perioperative period in civilians injured in the Syrian Civil War. Brazilian Journal of Anesthesiology (English Edition). 2015;65(6):445–9.

22. Arafat S, Alsabek MB, Ahmad M, Hamo I, Munder E. Penetrating abdominal injuries during the Syrian war: Patterns and factors affecting mortality rates. Injury. 2017;48(5):1054–7.

23. Çelikel A, Karbeyaz K, Kararslan B, Arslan MM, Zeren C. Childhood casualties during civil war: Syrian experience. Journal of forensic and legal medicine. 2015;34:1–4.

24. Aras M, Altaş M, Yılmaz A, Serarslan Y, Yılmaz N, Yengil E, et al. Being a neighbor to Syria: A retrospective analysis of patients brought to our clinic for cranial gunshot wounds in the Syrian civil war. Clinical neurology and neurosurgery. 2014;125:222–8.

25. Ozdogan HK, Karateke F, Ozdogan M, Cetinalp S, Ozyazici S, Gezercan Y, et al. The Syrian civil war: The experience of the Surgical Intensive Care Units. Pak J Med Sci. 2016;32(3):529–33.

26. Barhoum M, Tobias S, Elron M, Sharon A, Heija T, Soustiel JF. Syria civil war: Outcomes of humanitarian neurosurgical care provided to Syrian wounded refugees in Israel. Brain Inj. 2015;29(11):1370–5.

27. Guha-Sapir D, Schlüter B, Rodriguez-Llanes JM, Lillywhite L, Hicks MH-R. Patterns of civilian and child deaths due to war-related violence in Syria: a comparative analysis from the Violation Documentation Center dataset, 2011–16. The Lancet Global Health. 2018;6(1):e103–e10.

28. Er E, Çorbacıoğlu ŞK, Güler S, Aslan Ş, Seviner M, Aksel G, et al. Analyses of demographical and injury characteristics of adult and pediatric patients injured in Syrian civil war. Amer J Emerg Med. 2017;35(1):82–6.

29. Guha-Sapir D, Rodriguez-Llanes JM, Hicks MH, Donneau A-F, Coutts A, Lillywhite L, et al. Civilian deaths from weapons used in the Syrian conflict. BMJ : British Medical Journal. 2015;351:h4736.

30. Trelles M, Dominguez L, Tayler-Smith K, Kisswani K, Zerboni A, Vandenborre T, et al. Providing surgery in a war-torn context: the Médecins Sans Frontières experience in Syria. Conflict and Health. 2015;9(1):36.

31. Duramaz A, Bilgili, M.G., Bayram, B., et al. Orthopedic trauma surgery and hospital cost analysis in refugees; the effect of the Syrian civil War. International orthopaedics. 2017;41.

32. Institute for Health Metrics and Evaluation. How long do people live? - Syria Seattle [Available from: http://www.healthdata.org/syria.

33. Violations Documentation Center in Syria. Violations Documentation Center in Syria - Killed and Detained Dataset. Switzerland 2019.

34. Motaparthy P. Maybe We Live, Maybe We Die. New York: Human Rights Watch; 2014.

35. Geneva Call. Syria: new measures taken by the Kurdish People’s Protection Units to stop recruiting children under 18 Geneva: Geneva Call; 2018 [Available from: https://www.genevacall.org/syria-new-measures-taken-by-the-kurdish-peoples-protection-units-to-stop-using-children-under-18/.

36. UNHCR. Syria Regional Refugee Response - Inter-agency Information Sharing Portal Geneva: UNHCR; 2015 [Available from: http://data.unhcr.org/syrianrefugees/regional.php.

37. Mowafi H, Hariri M, Alnahhas H, Ludwig E, Allodami T, Mahameed B, et al. Results of a Nationwide Capacity Survey of Hospitals Providing Trauma Care in War-Affected Syria. JAMA Surg. 2016.

38. Spiegel PB, Salama P. War and mortality in Kosovo, 1998–99: an epidemiological testimony. Lancet. 2000;355(9222):2204–9.

39. Aboutanos MB, Baker SP. Wartime Civilian Injuries: Epidemiology and Intervention Strategies. Journal of Trauma and Acute Care Surgery. 1997;43(4):719–26.

40. Nerlander MP, Haweizy RM, Wahab MA, Älgå A, von Schreeb J. Epidemiology of Trauma Patients from the Mosul Offensive, 2016–2017: Results from a Dedicated Trauma Center in Erbil, Iraqi Kurdistan. World J Surg. 2019;43(2):368–73.

41. Assistance Coordination Unit. HeFRA SI-2. Gaziantep: Assistance Coordination Unit; 2015 April 2015.

42. Cluster UH. Health Resources Availability Mapping System (HeRAMS): Health Facilities Report. Gaziantep: UN Health Cluster; 2014.

43. Glance LG, Osler TM, Mukamel DB, Dick AW. Outcomes of adult trauma patients admitted to trauma centers in pennsylvania, 2000-2009. Archives of Surgery. 2012;147(8):732–7.

44. World Health Organization. Injury Surveillance Guidelines. Geneva: World Health Organization; 2001.

