## Appendix A for "Analysis of 193,618 trauma patient presentations in war-affected Syria from July 2013 to July 2015"

**Appendix A: Rule-based procedure to impute missing data elements**

| **Scenario** | **Missing Element** | **Action Taken** |
| --- | --- | --- |
| Procedure(s) coded routinely performed in an operating theatre and not commonly done at bedside (e.g. laparotomy, cranial surgery) | Emergency Department (ED) Disposition (ED Dispo) | Set ED Disposition to ADMIT |
| Procedure(s) coded minor surgical and routinely done at bedside (eg. Laceration repair, closed reduction of fracture) | Emergency Department (ED) Disposition (ED Dispo) | If Length of Stay ≥ 1 day then set Emergency Department Disposition to ADMIT  If Length of Stay ≤ 1 day or NOT RECORDED, then set ED Dispo to Discharged |
| Any patient with an ICU procedure code or with procedure code indicating mechanical ventilation | Emergency Department (ED) Disposition (ED Dispo) | Set ED Disposition to ICU ADMIT |
| Procedure code of mechanical ventilation | Procedure code missing for intubation | Emergency Department Disposition set to ICU ADMIT;  Endotracheal Intubation added to procedure code; |
| Procedure code for massive resuscitation | Emergency Department (ED) Disposition (ED Dispo) | Set ED Disposition to ICU ADMIT |
| Patient transferred to another facility for further treatment or definitive care | Time decision to transfer made | Convention for all transfers is that ED Disposition set to ADMIT and Hospital Disposition set to TRANSFER OUT;  Receiving hospital Emergency Department Disposition set to ADMIT or ICU ADMIT, as appropriate. The network of hospitals is not integrated enough to track patients reliably in the database. |
| Incomplete documentation of either injury, affected body part, mechanism, or procedure performed | Injury mechanism, body part, or Procedure code | If mechanism missing, then noted as INJURY;  If body part missing, then noted as BODY;  If procedure code missing but known to have undergone procedure (e.g. surgical outcome noted) , then PROCEDURE CODE noted as UNKNOWN SURGERY |
| Final disposition noted as dead | Missing discharge date, no procedure code recorded | Set ED Disposition to DEAD IN ED |
