## Appendix B for "Analysis of 193,618 trauma patient presentations in war-affected Syria from July 2013 to July 2015"

**Appendix Table B: Predicting Death Among Admitted Presentations Using Logistic Regression**

| **Variable** | **Female** | |  | **Male** | |  | **Combined** | |
| --- | --- | --- | --- | --- | --- | --- | --- | --- |
|  | **OR (95% CI)** | **adjusted OR (95% CI)** |  | **OR (95% CI)** | **adjusted OR (95% CI)** |  | **OR (95% CI)** | **adjusted OR (95% CI)** |
| **Gender** |  |  |  |  |  |  |  |  |
| Female |  |  |  |  |  |  | 1·35 (1·07-1·71) | 1·10 (0·98-1·25) |
| **Age Group** |  |  |  |  |  |  |  |  |
| 0 to 2 | 5·69 (3·82-8·49) | 4·45 (3·11-6·36) |  | 3·58 (1·87-6·88) | 2·21 (1·52-3·21) |  | 4·26 (2·44-7·46) | 2·57 (1·85-3·57) |
| 3 to 12 | 0·91 (0·59-1·41) | 1·66 (1·17-2·35) |  | 1·01 (0·80-1·29) | 0·97 (0·77-1·22) |  | 1·02 (0·78-1·32) | 1·06 (0·86-1·32) |
| 13 to 18 | 0·54 (0·25-1·16) | 1·02 (0·49-2·13) |  | 0·72 (0·52-1·00) | 0·72 (0·52-1·01) |  | 0·68 (0·47-0·99) | 0·76 (0·54-1·06) |
| 31 to 40 | 0·56 (0·30-1·03) | 1·08 (0·70-1·65) |  | 0·93 (0·72-1·20) | 0·93 (0·76-1·14) |  | 0·84 (0·62-1·15) | 0·94 (0·79-1·11) |
| 41 to 50 | 0·91 (0·64-1·29) | 1·53 (1·01-2·33) |  | 1·34 (1·01-1·78) | 1·20 (0·87-1·65) |  | 1·24 (0·96-1·60) | 1·21 (0·92-1·60) |
| 51 to 60 | 1·50 (1·08-2·08) | 2·22 (1·59-3·09) |  | 1·66 (1·25-2·21) | 1·49 (1·15-1·92) |  | 1·65 (1·35-2·03) | 1·55 (1·26-1·91) |
| 60+ | 3·59 (2·55-5·06) | 4·17 (2·78-6·25) |  | 2·46 (1·61-3·75) | 1·87 (1·34-2·62) |  | 2·99 (2·11-4·25) | 2·34 (1·79-3·05) |
| Unknown | 0·25 (0·12-0·54) | 0·47 (0·26-0·87) |  | 0·37 (0·19-0·73) | 0·38 (0·19-0·79) |  | 0·35 (0·18-0·68) | 0·39 (0·20-0·76) |
| **Injury** |  |  |  |  |  |  |  |  |
| Blast - Head | 4·01 (1·51-10·59) | 14·99 (4·29-52·38) |  | 7·29 (1·91-27·83) | 11·40 (2·87-45·36) |  | 6·64 (1·92-22·99) | 11·54 (3·18-41·95) |
| Blast - Chest | 0·41 (0·05-3·39) | 1·17 (0·13-10·83) |  | 1·30 (0·35-4·85) | 0·33 (0·08-1·38) |  | 1·16 (0·32-4·22) | 0·38 (0·10-1·51) |
| Blast - Abdomen | 0·14 (0·03-0·67) | 0·48 (0·07-3·27) |  | 1·91 (0·74-4·90) | 2·64 (0·96-7·26) |  | 1·51 (0·59-3·87) | 2·29 (0·84-6·26) |
| Blast - Spine | <0·01 (<0·01-<0·01) | <0·01 (<0·01-<0·01) |  | 78·68 (8·32-743·65) | 18·82 (0·83-427·56) |  | 49·35 (6·11-398·96) | 6·92 (0·14-336·87) |
| Blast - Extremities | 0·12 (0·01-0·96) | 0·17 (0·01-1·99) |  | 0·83 (0·27-2·57) | 0·86 (0·35-2·16) |  | 0·71 (0·23-2·20) | 0·75 (0·30-1·92) |
| Blunt - Head | 0·95 (0·43-2·11) | 1·98 (0·70-5·62) |  | 1·48 (0·66-3·29) | 2·62 (1·09-6·28) |  | 1·34 (0·62-2·91) | 2·38 (0·98-5·78) |
| Blunt - Chest | 7·72 (4·78-12·48) | 7·90 (3·94-15·84) |  | 5·69 (4·14-7·81) | 6·99 (4·09-11·97) |  | 6·47 (4·58-9·15) | 7·23 (4·16-12·57) |
| Blunt - Abdomen | 1·71 (1·17-2·51) | 3·21 (1·61-6·43) |  | 1·48 (0·95-2·31) | 2·57 (1·49-4·44) |  | 1·67 (1·18-2·37) | 2·80 (1·67-4·67) |
| Blunt - Spine | <0·01 (<0·01-<0·01) | <0·01 (<0·01-<0·01) |  | <0·01 (<0·01-<0·01) | <0·01 (<0·01-<0·01) |  | <0·01 (<0·01-<0·01) | <0·01 (<0·01-<0·01) |
| Blunt - Extremities | 0·07 (0·02-0·21) | 0·17 (0·06-0·47) |  | 0·14 (0·08-0·25) | 0·26 (0·16-0·44) |  | 0·12 (0·07-0·22) | 0·24 (0·16-0·37) |
| Burn - Head | 0·27 (0·06-1·20) | 0·60 (0·06-6·50) |  | 0·71 (0·29-1·72) | 0·73 (0·36-1·46) |  | 0·59 (0·30-1·15) | 0·73 (0·36-1·48) |
| Burn - Chest | <0·01 (<0·01-<0·01) | <0·01 (<0·01-<0·01) |  | 4·55 (1·16-17·83) | 3·91 (1·68-9·13) |  | 3·16 (0·80-12·52) | 2·77 (1·26-6·09) |
| Burn - Abdomen | 0·96 (0·42-2·16) | 5·69 (1·86-17·40) |  | 2·53 (0·56-11·49) | 1·19 (0·57-2·48) |  | 2·08 (0·57-7·61) | 1·83 (0·91-3·71) |
| Burn - Spine | <0·01 (<0·01-<0·01) | <0·01 (<0·01-<0·01) |  | 134·75 (11·34-1601·13) | 0·51 (0·02-14·78) |  | 63·41 (5·78-695·70) | 1·41 (0·04-54·26) |
| Burn - Extremities | 0·08 (0·01-0·58) | 0·22 (0·03-1·68) |  | 1·04 (0·33-3·28) | 0·89 (0·36-2·21) |  | 0·74 (0·24-2·31) | 0·72 (0·30-1·71) |
| Gunshot - Head | 1·04 (0·20-5·35) | 3·90 (0·83-18·41) |  | 3·46 (1·88-6·36) | 7·55 (3·64-15·68) |  | 3·03 (1·58-5·82) | 7·27 (3·49-15·12) |
| Gunshot - Chest | 1·27 (0·42-3·84) | 4·20 (1·69-10·43) |  | 1·41 (0·74-2·67) | 3·32 (1·87-5·91) |  | 1·35 (0·68-2·69) | 3·41 (1·92-6·08) |
| Gunshot - Abdomen | 1·06 (0·39-2·93) | 3·90 (1·79-8·47) |  | 0·86 (0·46-1·63) | 1·80 (1·03-3·14) |  | 0·88 (0·44-1·77) | 2·02 (1·16-3·50) |
| Gunshot - Spine | <0·01 (<0·01-<0·01) | <0·01 (<0·01-<0·01) |  | 1·35 (0·43-4·25) | 3·75 (1·03-13·68) |  | 1·11 (0·36-3·37) | 3·14 (0·90-10·92) |
| Gunshot - Extremities | 0·14 (0·04-0·56) | 0·48 (0·22-1·04) |  | 0·11 (0·05-0·22) | 0·26 (0·16-0·43) |  | 0·11 (0·05-0·24) | 0·28 (0·17-0·47) |
| Shrapnel - Head | 2·44 (1·16-5·14) | 5·94 (2·78-12·68) |  | 3·72 (2·08-6·67) | 5·94 (3·06-11·51) |  | 3·41 (1·85-6·27) | 5·94 (3·08-11·43) |
| Shrapnel - Chest | 1·79 (0·83-3·88) | 4·15 (2·40-7·18) |  | 2·17 (1·23-3·83) | 3·15 (2·00-4·97) |  | 2·07 (1·13-3·78) | 3·27 (2·11-5·07) |
| Shrapnel - Abdomen | 1·27 (0·61-2·67) | 3·49 (1·80-6·75) |  | 1·89 (1·41-2·53) | 3·15 (2·17-4·56) |  | 1·74 (1·23-2·47) | 3·18 (2·19-4·62) |
| Shrapnel - Spine | 1·04 (0·20-5·40) | 3·67 (0·71-18·88) |  | 3·50 (1·40-8·76) | 4·07 (2·04-8·11) |  | 3·00 (1·24-7·26) | 3·87 (1·99-7·55) |
| Shrapnel - Extremities | 0·29 (0·13-0·66) | 0·84 (0·42-1·69) |  | 0·37 (0·20-0·68) | 0·77 (0·44-1·34) |  | 0·35 (0·18-0·67) | 0·78 (0·45-1·33) |
| Other - Head | <0·01 (<0·01-<0·01) | <0·01 (<0·01-<0·01) |  | 0·11 (0·01-0·88) | 0·16 (0·02-1·47) |  | 0·09 (0·01-0·71) | 0·14 (0·02-1·27) |
| Other - Chest | <0·01 (<0·01-<0·01) | <0·01 (<0·01-<0·01) |  | 0·50 (0·12-2·07) | 1·31 (0·36-4·83) |  | 0·38 (0·10-1·46) | 1·04 (0·32-3·39) |
| Other - Abdomen | <0·01 (<0·01-<0·01) | <0·01 (<0·01-<0·01) |  | 0·24 (0·06-1·02) | 0·54 (0·13-2·18) |  | 0·18 (0·04-0·77) | 0·43 (0·11-1·74) |
| Other - Spine | NA (NA-NA) |  |  | <0·01 (<0·01-<0·01) | <0·01 (<0·01-<0·01) |  | <0·01 (<0·01-<0·01) | <0·01 (<0·01-<0·01) |
| Other - Extremities | <0·01 (<0·01-<0·01) | <0·01 (<0·01-<0·01) |  | <0·01 (<0·01-<0·01) | <0·01 (<0·01-<0·01) |  | <0·01 (<0·01-<0·01) | <0·01 (<0·01-<0·01) |

N = 10,557 female; 48,830 male; 57,625 overall. 95% confidence intervals are estimated using cluster-robust standard errors clustering on facility.

Adjusted OR (aOR) were adjusted for all the other variables in the model including age, gender, and injury mechanism-body part combination
